# High blood uric acid is associated with reduced risks of mild cognitive impairment among older adults in China: a 9-year prospective cohort study

**DOI:** 10.1101/2021.07.24.21261062

**Authors:** Chen Chen, Xueqin Li, Yuebin Lv, Zhaoxue Yin, Feng Zhao, Yingchun Liu, Chengcheng Li, Saisai Ji, Jinhui Zhou, Yuan Wei, Xingqi Cao, Jiaonan Wang, Heng Gu, Feng Lu, Zuyun Liu, Xiaoming Shi

## Abstract

**Background:** It remains unsolved that whether blood uric acid (UA) is a neuroprotective or neurotoxic agent. This study aimed to evaluate the longitudinal association of blood UA with mild cognitive impairment (MCI) among older adults in China.

**Methods:** A total of 3103 older adults (aged 65+ years) free of MCI at baseline were included from the Healthy Aging and Biomarkers Cohort Study (HABCS). Blood UA level was determined by the uricase colorimetry assay and analyzed as categorical (by quartile) variables. Global cognition was assessed using the Mini-Mental State Examination four times between 2008 and 2017, with a score below 24 being considered as MCI. Cox proportional hazards models were used to examine the associations.

**Results:** During a 9-year follow-up, 486 (15.7%) participants developed MCI. After adjustment for all covariates, higher UA had a dose-response association with a lower risk of MCI (all *P* _for trend_< 0.05). Participants in the highest UA quartile group had a reduced risk (hazard ratio [HR], 0.73; 95% [CI]: 0.55-0.96) of MCI, compared with those in the lowest quartile group. The associations were still robust even when considering death as a competing risk. Subgroup analyses revealed that these associations were statistically significant in younger older adults (65-79 years) and those without hyperuricemia.

**Conclusions:** High blood UA level is associated with reduced risks of MCI among Chinese older adults, highlighting the potential of managing UA in daily life for maintaining late-life cognition.

## Background

Uric acid (UA) is a chemical which is created when the human body breaks down purines and then catalyzed by xanthine oxidase. Blood UA level is determined by the residual quantities between dietary purine intake and renal excretion. Hyperuricemia, the status of an abnormally high level of UA in the blood, is a prerequisite for gout (1). Both hyperuricemia and gout confer a high risk for subsequent metabolic syndrome (2) and cardiovascular diseases (CVDs) (3, 4). However, as a natural antioxidant, the antioxidant properties of UA may protect against the detrimental effect of oxidative stress among people with central nervous system disorders (5, 6). Some debates have been raised that urate-lowering-therapies might paradoxically expose patients to a high risk of neurodegenerative disease (such as dementia) (7, 8).

To date, many studies have explored the associations between blood UA levels and cognition or dementia in developed countries, but the results remain in dispute. Positive (5, 9-15), negative (16-18), U-shaped (19), or no associations (20) of blood UA levels with cognition have been reported. In China, some cross-sectional studies support the neuroprotective role of UA in cognition. However, most of them were limited to their cross-sectional nature and the relatively small sample size (21-26). To our best knowledge, only three relevant longitudinal studies have been conducted, with one on Parkinson’s disease (PD) (27) or one on dementia (28). The only one on cognition was limited to its relatively short follow-up time (ranged from 1.3 to 2.4 years) (29) (For detailed information see Supplementary Table S1). Therefore, investigations on the long-term effect of UA on cognition in general Chinese populations are urgently needed.

Therefore, based on 9-year follow-up data from a community-based multi-wave cohort of older adults (65 years and older), the Healthy Aging and Biomarkers Cohort Study (HABCS), this study aimed to evaluate the longitudinal associations of blood UA levels with the risks of incident mild cognitive impairment (MCI) in general Chinese older population.

## Methods

### Study population

The HABCS was launched in eight longevity areas in China in 2008, collecting comprehensive data, such as health status, and behavioral, laboratory, and anthropometric measurements. The follow-up surveys were conducted in 2012, 2014, and 2017. Due to the high death rate in older adults, new participants were recruited in the follow-up surveys to maintain a stable sample size for the dynamic cohort. Details of this survey design have been described elsewhere (30). A total of 5074 older adults from the 2008 survey and the recruits from the follow-up surveys (2012-2014 waves) from the HABCS were included in this study. We excluded 606 participants aged <65 years, 6 participants with missing data on sex, 368 participants on Mini-Mental State Examination (MMSE) score, and 195 participants on UA levels, and then excluded 796 participants with prevalent CI at baseline, leaving 3103 participants free of CI for the final longitudinal analysis (Fig. 1). All participants or their legal representatives gave written informed consent to participate in the baseline and follow-up surveys. The HABCS was approved by the ethics committee of Peking University and Duke University.

**Fig. 1.**
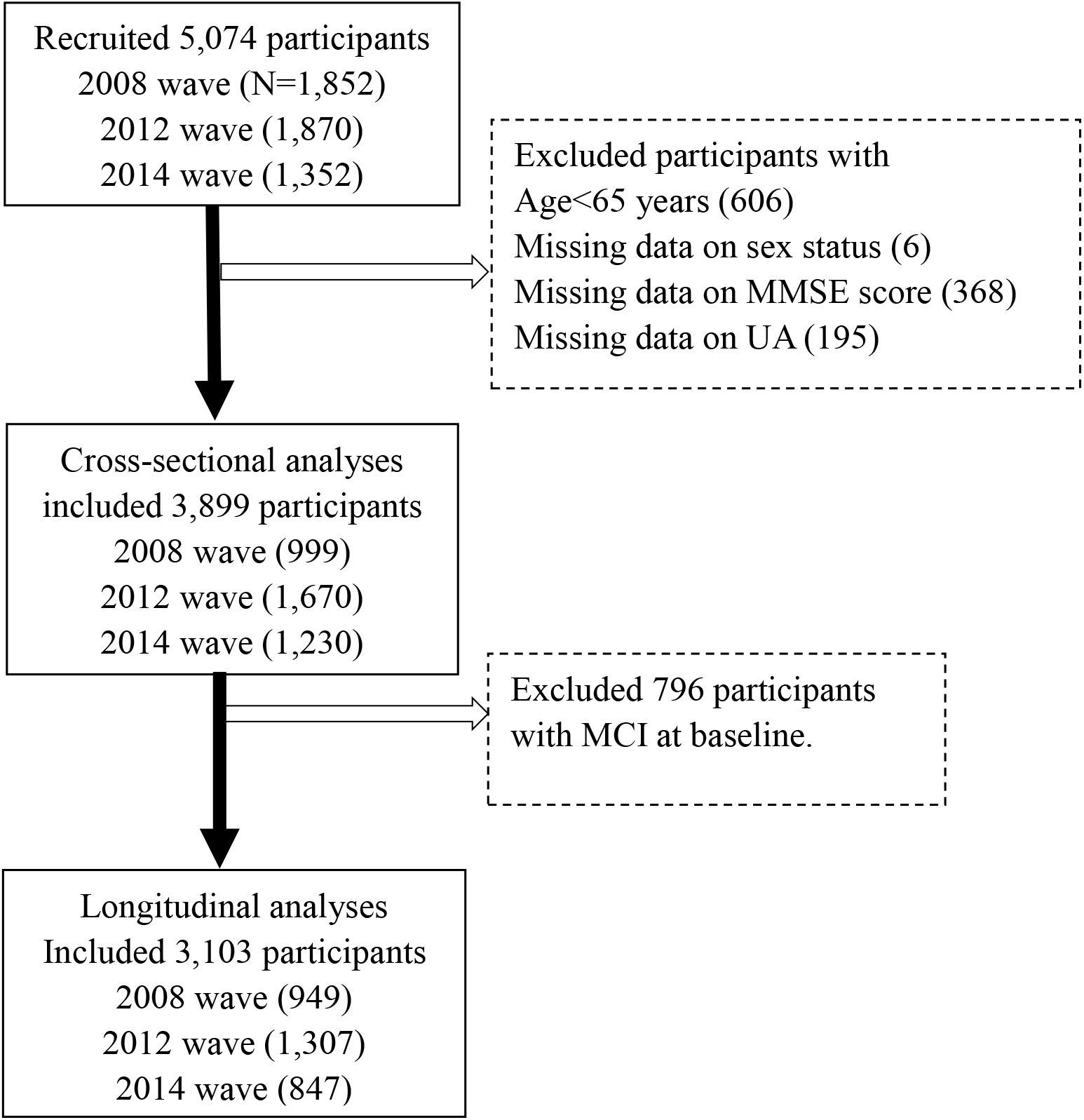
Flowchart of study participant enrollment in this study. Abbreviations: MMSE, Mini-Mental State Examination. UA, uric acid.

### Assessment of blood UA levels

Fastening venous blood was collected from all participants in the baseline wave (the year 2008-2014). The plasma was separated and stored at −20 °C and delivered to the laboratory at Capital Medical University in Beijing for unified detection. Blood UA level was determined by uricase colorimetry assay. Details on quality control and assessments in the laboratory were described elsewhere (31). Blood UA levels >420 μmol/L in men and UA levels >360 μmol/L in women were defined as hyperuricemia (32), which indicates above the UA normal range. Sex-specific quartiles of UA were constructed and used in the analysis.

### Assessment of cognition

Cognition was assessed in the years 2008, 2012, 2014, and 2017, using the modified version of the widely used MMSE questionnaire (33). The MMSE included 6 cognitive domains (orientation, working memory, concentration, memory recall, language, and visuospatial ability), with a total score ranging from 0 to 30, and the higher score indicates better cognition. We categorized cognition into two levels: no MCI (24≤ MMSE ≤ 30) and MCI (MMSE<24) as done previously (34). We set MCI as the outcome and calculated the follow-up time for each participant from baseline to either the date of incident MCI, loss to follow-up (including death), or the end of the study period, whichever came first.

### Covariates

Age, sex, education, current marital status, current smoking, current alcohol consuming, regular exercise, and adequate medical service were considered. Education was categorized as having more than 1 year of schooling or no formal education. Current marital status was categorized as currently married or others. Current smoking and current alcohol consumption were categorized as with or without status. Regular exercise was classified into “yes” or “no” by the question “Do you do exercises regularly at present, including walking, playing ball, running, and Qigong?” Hypertension was defined as yes if a participant had an SBP ⩾140 mmHg and/or a DBP ⩾90, or self-reported suffering from hypertension. Diabetes mellitus was defined as yes if a participant had fasting glucose ⩾ 7.0 mmol/L or self-reported current diabetes medication use. Self-reported history of heart disease, stroke and CVD were also considered. Standardized protocols were used to collect measurements of weight, height, and waist circumference (WC) (35). Central obesity was defined as 85 cm or larger of WC in men and 80 cm or larger of WC in women. These covariates have been demonstrated as important correlations with blood UA level (36), or cognition (37), or both (30).

### Statistical analyses

Descriptive statistics of the baseline characteristics were presented as means± standard deviation (SD) or percentages among the full sample and by the sex-specific UA quartile groups. The Cox proportional hazards model was used to calculate the hazard ratios (HRs) and 95% confidence intervals (CIs) for the association between blood UA quartiles (the lowest UA quartile as a reference category) and the risk of MCI. Three models were considered. In model 1, we included age, sex, and education. In model 2, we further included current smoking, current alcohol consuming, marital status, regular exercise, BMI, central obesity, and adequate medical service based on model 1. In model 3, we additionally adjusted for hypertension, diabetes mellitus, heart disease, and stroke and CVD based on model 2. Then, given the high proportion of death in older adults, we used Fine & Gray competing risk models to re-examine the association between blood UA and risk of developing MCI, using the similar models as above. Additionally, we used restricted cubic splines (RCS) to flexibly model and visualize the dose-response association of blood UA levels with the risk of MCI with three sex-specific knots at the 5th, 50th, and 95th percentiles of blood UA distribution in model 1.

We performed subgroup analyses of the association between blood UA and the risk of developing MCI by age (65-79 vs ⩾ 80 years), sex (men vs women), and hyperuricemia (without hyperuricemia vs with hyperuricemia) using model 3. We added the interaction items (e.g., age group * UA quantiles) to determine whether the associations differed by the subgroup.

A series of sensitivity analyses were conducted to verify the robustness of the above estimations. First, we compared the baseline characteristics of the included and excluded participants. Second, because we included new participants enrolled in the follow-up surveys, we also compared the baseline characteristics of study participants by different enrolled times. Third, we evaluated the associations and dose-response relationship between blood UA and baseline cognition score, to confirm the findings from previous studies using a cross-section design.

A *P*-value of less than 0.05 (two-tailed) was considered. All analyses were carried out in SAS version 9.4 (SAS Institute, Cary, NC).

## Results

### Basic characteristics of study participants

The mean (SD) age of 3103 older adults was 85.1 (±11.7) years, and approximately 54% (n=1,688) were women. Participants with higher blood UA were more likely to have traditional cardiovascular disease risk factors (drinking, central obesity, hypertension and diabetes mellitus), while had higher proportion of the assumed healthy behaviors (regular exercise) (all *P*<0.05, Table 1).

**Table 1.** Baseline characteristics of study participants by UA quartiles ^a^

| Characteristics | Total<br>(N=3103) | Q<sub>1</sub> (N=773) | Q<sub>2</sub> (N=777) | Q<sub>3</sub> (N=776) | Q<sub>4</sub> (N=777) | P Value |
| --- | --- | --- | --- | --- | --- | --- |
| Age, mean ±SD, years | 85.1 ± 11.7 | 82.8 ± 12.2 | 84.8 ± 11.6 | 85.8 ± 11.4 | 86.8 ± 11.2 | <0.001 |
| Sex, women (%) | 1688 (54.4) | 422 (54.6) | 421 (54.2) | 424 (54.6) | 421 (54.2) | 0.996 |
| ≥ years of education,<br>yes (%) | 1098 (36.3) | 274 (36.2) | 258 (33.9) | 276 (36.6) | 290 (38.6) | 0.319 |
| Currently married, yes<br>(%) | 1170 (38.5) | 318 (41.6) | 294 (38.6) | 292 (38.4) | 266 (35.5) | 0.113 |
| Regular exercise, yes (%) | 552 (18.4) | 105 (13.9) | 150 (19.7) | 144 (19.0) | 153 (20.8) | 0.003 |
| Current smoking, yes (%) | 573 (18.8) | 152 (19.9) | 134 (17.4) | 142 (18.5) | 145 (19.3) | 0.628 |
| Current alcohol drinking,<br>yes (%) | 510 (16.8) | 135 (17.6) | 101 (13.3) | 139 (18.2) | 135 (18.0) | 0.028 |
| BMI, mean ±SD, kg/m<sup>2</sup> | 21.5 ± 10.1 | 20.8 ± 3.4 | 21.9 ± 17.2 | 21.4 ± 6.3 | 21.8 ± 7.7 | 0.170 |
| Central obesity <sup>b</sup>, yes (%) | 1303 (42.3) | 308 (40.1) | 315 (41.0) | 318 (41.3) | 362 (46.8) | 0.031 |
| Adequate medical<br>service, yes (%) | 2864 (93.9) | 706 (92.4) | 722 (94.1) | 728 (94.9) | 708 (94.1) | 0.212 |
| Hypertension, yes (%) | 1730 (55.8) | 352 (45.6) | 410 (52.9) | 473 (61.0) | 495 (63.8) | <0.001 |
| Diabetes mellitus, yes<br>(%) | 331 (10.7) | 74 (9.6) | 65 (8.4) | 99 (12.8) | 93 (12.0) | 0.017 |
| Heart Disease, yes (%) | 232 (7.7) | 58 (7.7) | 46 (6.1) | 61 (8.1) | 67 (8.9) | 0.212 |
| Stroke/CVD, yes (%) | 165 (5.4) | 50 (6.6) | 35 (4.6) | 35 (4.6) | 45 (5.9) | 0.232 |

Abbreviations: UA, uric acid; SD, standard deviation; BMI, body mass index; CVD, cardiovascular disease.
Note: Values are given as No. (%) unless otherwise stated.
a The cutoff values were 213.9, 265.0, and 320.9 μmol/L for women, and 252.0, 305.6, and 365.1 μmol/L for men.
b Central obesity was defined as waist circumference ≥ 80 cm in women and waist circumference ≥ 85 cm in men.

### Associations of blood UA with the incidence of MCI

During a 9-year follow-up, 486 (15.7%) participants developed MCI. From three adjusted models in Table 2, we found that higher UA levels had a lower risk of developing MCI. After full adjustment of covariates in model 3, compared with participants in the lowest quartile group of UA level (Q1), those in the highest quartile group (Q4) had a 27% lower risk of MCI [HR_Q4 vs Q1_=0.73 (95% CIs: 0.55-0.96)]. The association was still robust after considering death as a competing risk [HR_Q4 vs Q1_=0.67 (95% CIs: 0.51-0.88)]. The dose-response relationship between blood UA and the risk of MCI was shown in Fig. 2 (*P* _for non-linear_=0.080).

**Table 2.** Longitudinal associations of blood UA levels with the risk of MCI.

| **Blood UA**<br>**($\mu\text{mol/L}^a$)** | **Model 1**<br>**HR (95% CI)** | **Model 2**<br>**HR (95% CI)** | **Model 3**<br>**HR (95% CI)** |
| --- | --- | --- | --- |
| Cox proportional hazards model |  |  |  |
| Q1 | 1.00 [Reference] | 1.00 [Reference] | 1.00 [Reference] |
| Q2 | 0.84 (0.66, 1.06) | 0.84 (0.66, 1.07) | 0.82 (0.64, 1.06) |
| Q3 | 0.81 (0.64, 1.04) | 0.83 (0.65, 1.06) | 0.73 (0.56, 0.95) |
| Q4 | 0.74 (0.57, 0.96) | 0.75 (0.57, 0.98) | 0.73 (0.55, 0.96) |
| P<sub>for trend</sub> | 0.022 | 0.036 | 0.013 |
| Fine & Gray competing risks model<sup>b</sup> |  |  |  |
| Q1 | 1.00 [Reference] | 1.00 [Reference] | 1.00 [Reference] |
| Q2 | 0.90 (0.71, 1.14) | 0.89 (0.69, 1.14) | 0.88 (0.68, 1.13) |
| Q3 | 0.89 (0.70, 1.13) | 0.80 (0.62, 1.04) | 0.79 (0.61, 1.02) |
| Q4 | 0.70 (0.54, 0.91) | 0.67 (0.51, 0.88) | 0.67 (0.51, 0.88) |
| P<sub>for trend</sub> | 0.014 | 0.018 | 0.003 |

Abbreviations: UA, uric acid; HR, hazard, ratio; CI, confidence interval.
Model 1 adjusted for age, sex and education; model 2 additionally adjusted for drinking, smoking, marital status, regular exercise, body mass index, central obesity, adequate medical service; model3 additionally adjusted for hypertension, diabetes mellitus, self-reported history of heart disease, and stroke and cardiovascular disease.
a The cutoff values were 213.9, 265.0, and 320.9 $\mu\text{mol/L}$ for women, and 252.0, 305.6, and 365.1 $\mu\text{mol/L}$ for men.
b Adjusted for death as competing risk.

**Fig. 2.**
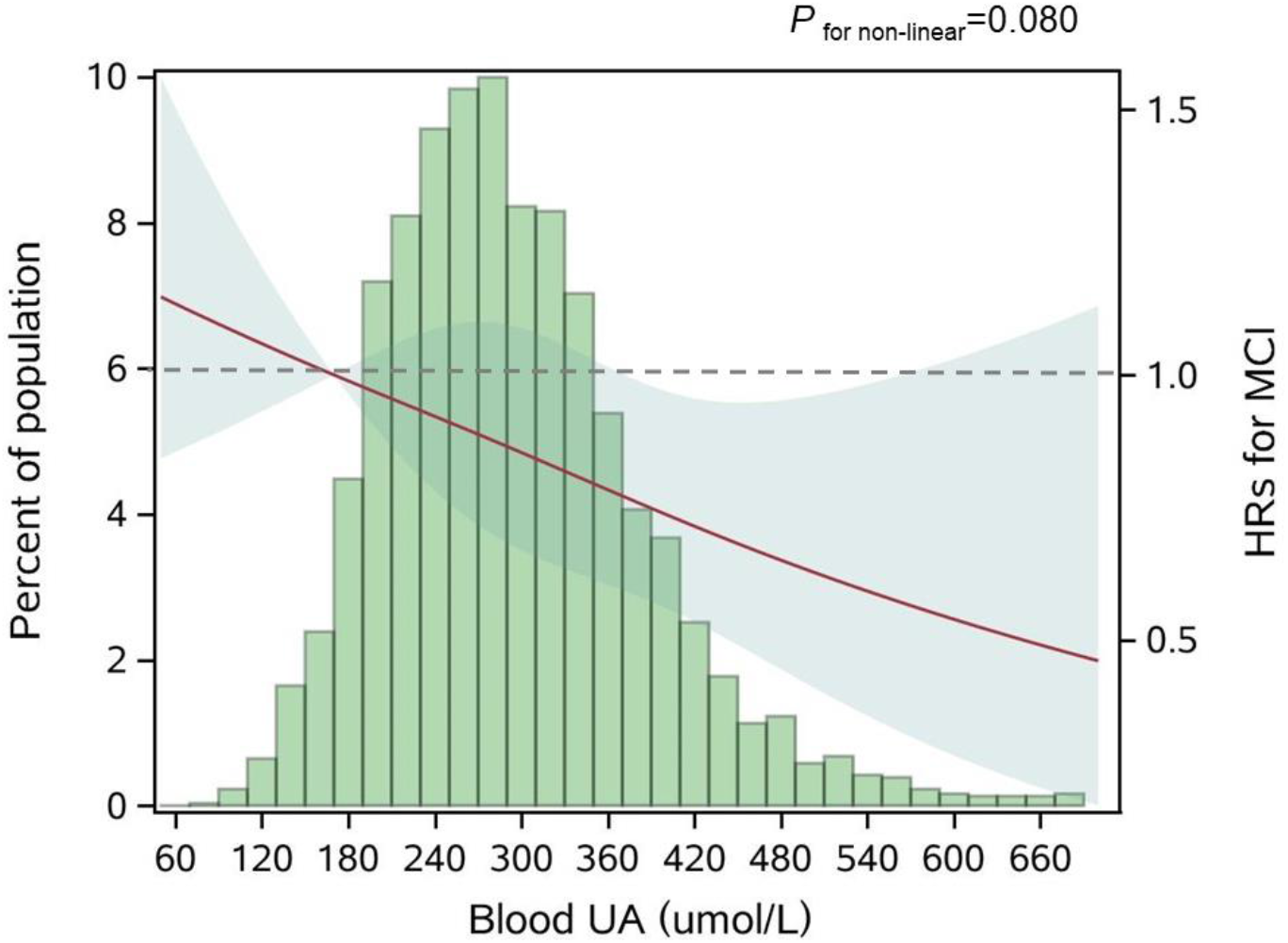
Adjusted dose-response association between blood UA and risk for MCI. Abbreviations: UA, uric acid; HR, hazard ratio; MCI, mild cognitive impairment. Note: Blood UA was coded using a restricted cubic spline (RCS) function with three sex-specific knots, which approximately corresponded to the 5th (170.0 μmol/L), 50th (281.6 μmol/L), and 95th (454.3 μmol/L) percentiles of blood UA distribution. Solid red line represents the adjusted hazard ratio for the risk of MCI for any value of UA compared to participants with 170.0 μmol/L (P5) of blood UA level, with light green shaded areas showing 95% confidence intervals derived from restricted cubic spline regressions. The green histograms show the fraction of the population with the different levels of blood UA. Dashed gray line refers to the reference for the association at a HR of 1.0. P for non-linear= 0.080.

### Subgroup analyses

Subgroup analyses stratified by age and sex are shown in Fig. 3. The associations were more pronounced in the younger older adults (65-79 years) (*P* _for interaction_=0.034). Compared with the lowest quartile group of UA level (Q1), those in the highest quartile group (Q4) had a 50% lower risk of developing MCI [HR_Q4 vs Q1_=0.50 (95% CI: 0.26-0.98)]. Partially due to the relatively small sample size, some of the associations of blood UA levels with the risk of MCI became non-significant (e.g., that in women). In addition, we observed that the association of blood UA on the risk of MCI remained statistically significant among those without hyperuricemia [HR _Q4 vs Q1_=0.64 (95% CI: 0.48-0.85)] (Supplementary Table S2). However, no significant associations were found for older adults with UA levels above the normal range (i.e., with hyperuricemia) [HR_Q4 vs Q1_=2.19 (0.81-5.88)].

**Fig. 3.**
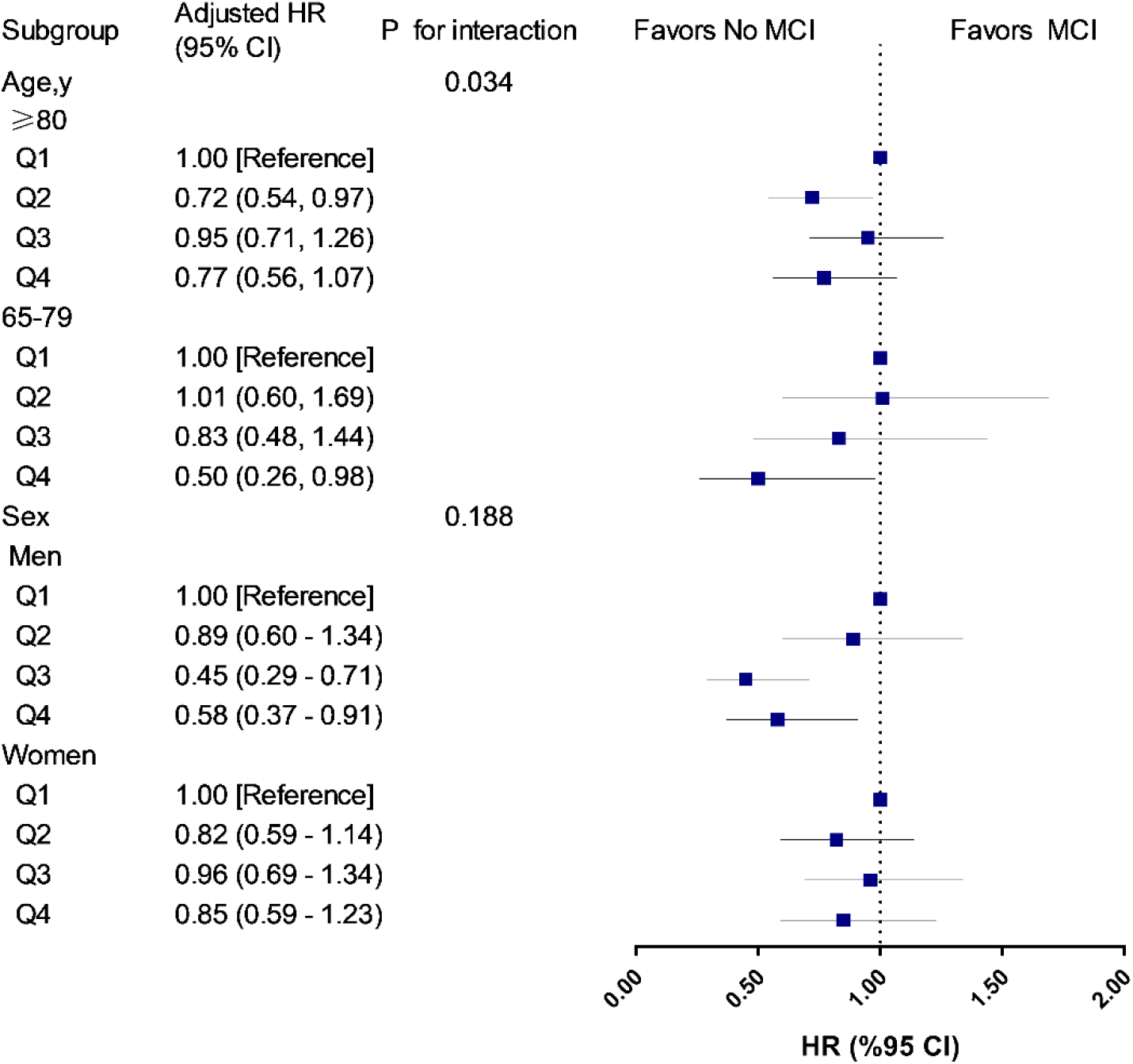
Associations of UA quartiles with the risk of MCI in age and sex subgroups. Abbreviations: HR, hazard ratio; CIs, confidence intervals. Note: The model was adjusted for age (not in age subgroup) and sex (not in sex subgroup), education, drinking, smoking, marital status, regular exercise, body mass index, central obesity, adequate medical service, hypertension, diabetes mellitus, self-reported history of heart disease, and stroke/CVD. Q1 was defined as the reference group. In the age subgroup, participants aged 65-79 years, the cutoff values of UA quartiles were 245.4, 291.6, and 355.4 μmol/L for men, and 195.2, 242.0, and 296.4 µmol/L for women; participants aged 80 years and older, the cutoff values of UA quartiles were 262.0, 315.0, and 377.1 μmol/L for men, and 220.0, 270.7, and 329.8 µmol/L for women. In the sex subgroup, the cutoff values were 252.0, 305.6, and 365.1 µmol/L for men, and 213.9, 265.0, and 320.9 µmol/L for women.

### Sensitivity analyses

In sensitivity analyses: (1) Differences in demographic characteristics were found between eligible and excluded participants. Those who were excluded were more likely to be younger and women, and had lower blood UA levels; and had more healthy characteristics (a higher proportion of education, and a lower proportion of smoking, central obesity, hypertension, and heart disease) (all P <0.05, Supplementary Table S3). (2) Different characteristics of participants by enrolled time (i.e., 2008 wave, 2012 wave, and 2014 wave) were observed (Supplementary Table S4). (3) Consistent results of higher UA levels associated with higher cognition scores were found using general linear models and dose-response testing (*P* _for non-linear_ =0.041) among 3899 participants in the cross-sectional analysis (Supplementary Table S5-S6 and Fig. S1).

## Discussion

In this community-based longitudinal study of 3103 Chinese older adults free of MCI at baseline, we found that higher UA was associated with a lower risk of developing MCI during the 9 years follow-up. Furthermore, these associations were generally stronger in the younger older adults (65-79 years) and those without hyperuricemia. The findings confirm the neuroprotective effect of UA in Chinese older populations.

The neuroprotective effects of blood UA we observed are generally consistent with that from the previous epidemiological studies. A recent cross-sectional study using data from Beijing Longitudinal Study on Aging II showed the beneficial role of high blood UA levels on CI (22). A cohort study from Netherland, with over 11-year follow-up, found that higher serum UA levels at baseline predicted better cognition in later life (5). Similarly, another cohort (13) identified the potential neuroprotective role of UA on Alzheimer’s disease in a 5-year follow-up. However, there are still some studies that failed to observe the neuroprotective role of UA in dementia (16-18). The heterogeneity in study populations and outcome measurements might partially explain the different findings across studies.

Several possible biological mechanisms may elucidate the observed association of higher normal UA levels with a lower risk of MCI in the current study. First, in a urate oxidase mouse model with hemi-parkinsonism, genic mutated urate oxidase resulted in increased brain urate concentrations and substantially attenuated toxic effects of 6-hydroxydopamine on neurochemicals and rotational behavior. Additionally, transgenic urate oxidase decreased brain urate, which exacerbated neural functional lesions (38). Second, UA exerts its important antioxidant property in human bodies by acting as a direct scavenger of oxygen and hydroperoxyl radicals and forming stable complexes with iron ions (39), which further reduces the risk of MCI (40, 41). Third, nutrition status might partly play a role in the UA-cognition relationship. Hypouricemia, a status of the abnormally low level of UA in the blood, is a well-established biomarker of poor nutritional status (42). Meanwhile, malnutrition is a serious issue in older adults with MCI or dementia (43).

In our study, the association of blood UA with the risk of MCI appeared to be more pronounced in the younger older adults (65-79 years) and those without hyperuricemia. The age-specific protective effects of high blood UA on the risk of MCI might be explained by the fact that the younger older adults have a lower aging rate, and to be more sensitive to the beneficial effects of blood UA on cognition (44). Furthermore, survivor bias might exist in our study population. The unexpected non-significant result among those without hyperuricemia might be due to occasional chance and the relatively small sample size (N _hyperuricemia_=404). Besides, from the dose-response test (Fig. 2), we noticed that the 95% CIs for the risk estimates became extremely wider when the UA levels were higher than about 420 μmol/L, which was around the cutoff value of the UA normal range (blood UA levels >420 μmol/L in men and UA levels >360 μmol/L in women were defined as hyperuricemia). These older adults with hyperuricemia might not share the same underlying etiology pathway relative to those without hyperuricemia.

The strengths of this study include its prospective cohort design and a relatively large sample size of older adults. However, several limitations should be considered. First, comprehensive neuropsychological assessments and the diagnoses of specific dementia or Parkinson’s disease were not available in our study, so we could not capture the detailed aspects and clinical stage of cognition. Nevertheless, i) we repeated measured the MMSE four times, which would allow us to capture the global patterns of cognitive fluctuation over time; ii) the MMSE was commonly used as part of the evaluation for possible dementia (34). Second, the UA levels were measured only once, and may not reflect the true value and ignore the fluctuation. Third, residual confounding could not be ignored, such as, we did not collect information on dietary habits and medication, which might serve as confounding factors in the UA-cognition association. Due to these limitations, we need to be cautious when interpreting our results. Given that our study focused on the Chinese older adults, more studies are warranted to further investigate the associations in other ethnic groups.

## Conclusion

By evaluating the associations of blood UA with the risk of MCI, our results suggested the protective role of high blood UA among Chinese older adults. The findings highlight the potential of managing UA in daily life for maintaining late-life cognition, with important clinical practice implications, and deserves further verifications.

## Acknowledgments

We appreciate all participants who attended the Healthy Aging and Biomarkers Cohort Study (HABCS).

## Data availability statement

The data that support the findings of this study are available from the corresponding author, Dr. Shi, upon reasonable request.

## Authors’ contributions

Study concept and design: Zuyun Liu, Xiaoming Shi; Acquisition, analysis, and interpretation of data: Chen Chen, Xueqin Li; Drafting of the manuscript: Chen Chen, Xueqin Li; Critical revision of the manuscript for important intellectual content: Yuebin Lv, Xingqi Cao, Zuyun Liu, Xiaoming Shi; Statistical analysis: Chen Chen, Xueqin Li; Obtained funding: Zuyun Liu, Xiaoming Shi; Administrative, technical, or material support: Zhaoxue Yin, Feng Zhao, Yingchun Liu, Chengcheng Li, Saisai Ji, Jinhui Zhou, Yuan Wei, Xingqi Cao, Jiaonan Wang, Heng Gu, Feng Lu. Supervision: Zuyun Liu, Xiaoming Shi.

