## Supplemental tables and figure for "High blood uric acid is associated with reduced risks of mild cognitive impairment among older adults in China: a 9-year prospective cohort study" (from another version of this preprint: version 1, 2021-07-28)

**Supplement information**

**Table S1.** Summary of previous studies on associations of blood UA with cognition and relevant outcomes

**Table S2.** Longitudinal associations of blood UA levels with the risk of MCI among participants without and with hyperuricemia.

**Table S3.** Baseline characteristics of the included and excluded study participants.

**Table S4.** Baseline characteristics of study participants by different enrolled times.

**Table S5.** Baseline characteristics of study participants by UA quartiles for the cross-sectional analyses

**Table S6.** Cross-sectional associations of blood UA levels with the cognitive score.

**Fig. S1.** Adjusted dose-response association between blood UA and cognition score at baseline.

**Table S1.** Summary of previous studies on associations of blood UA with cognition and relevant outcomes

| **First author** | **Year** | **Country** | **N** | **Age** | **Study types** | **CI** | **Dementia** | **PD** | **AD** |
| --- | --- | --- | --- | --- | --- | --- | --- | --- | --- |
| S. M. Euser [5] | 2009 | Netherlands | 4618 | ≥ 55 years | 11.1 years longitudinal | ↑ | ↑ |  |  |
| Michael C Irizarry [9] | 2009 | USA | 747 | mean: 73 years | 3 years longitudinal |  |  | ↑ |  |
| Busra Tuven [10] | 2017 | Turkey | 1347 | mean:76.72 years | cross-sectional | ↑ |  |  |  |
| Mary De Vera [11] | 2008 | Canada | 67 457 | ≥ 65 years | 8 years longitudinal |  |  | ↑ |  |
| Arrigo F Cicero [12] | 2014 | Italy | 288 | mean: 69 ± 6 years | cross-sectional | ↑ |  |  |  |
| Na Lu [13] | 2015 | UK | 298 029 | mean: 65 years | 5 years longitudinal |  |  |  | ↑ |
| Maria Pellecchia [14] | 2016 | Italy. | 70 | mean: 59 years | 4 years longitudinal | ↑ |  |  |  |
| B S Ye [15] | 2016 | US | 1064 | mean: 73.7 years | 2.9 years longitudinal | ↑ |  |  |  |
| Aug Latourte [16] | 2018 | French | 4931 | ≥ 65 years | 12 years longitudinal |  | ↓ |  |  |
| Jas A Singh [17] | 2018 | US | 1.7million | mean: 75.2 years | 6 years longitudinal |  | ↓ |  |  |
| Aniqa B Alam [18] | 2020 | USA | 11 169 | 47–70 years | 24.1 years longitudinal | ↓ | → |  |  |
| Rong Huang [19] | 2019 | China | 352 | ≥ 58 years | cross-sectional | U |  |  |  |
| Yuzhen Xu [21] | 2017 | China | 208 | ≥ 67 years | cross-sectional | ↑ |  |  |  |
| Shuangling Xiu[22] | 2017 | China | 10 039 | ≥ 55 years | cross-sectional | ↑ |  |  |  |
| Feng Wang [23] | 2017 | China | 1 866 | ≥ 60 years | cross-sectional | ↑ |  |  |  |
| Miao Liu [24] | 2017 | China | 2 102 | mean: 71.2 years | cross-sectional | ↑ |  |  |  |
| Yili Wu [25] | 2013 | China | 2 006 | 50–74 years | cross-sectional | ↑ |  |  |  |
| Jun Li [26] | 2010 | China | 694 | 90–108 years | cross-sectional | ↑ |  |  |  |
| Li-Yu Hu [27] | 2020 | China | 15 800 | median: 50 years | 13.36 years longitudinal |  |  | → |  |
| Jen-Yee Hong [28] | 2015 | China | 30 316 | ≥ 50 years | 4 years longitudinal |  | ↑ |  |  |
| Tong Wang [29] | 2017 | China | 12 798 | ≥ 45 years | 1.33-2.42 years longitudinal | ↑ |  |  |  |

Abbreviations: CI, cognitive impairment; AD, Alzheimer’s disease; PD, Parkinson's disease;

↑, refers to that UA was positive factor for cognition; ↓, refers to that UA was negative factor for cognition; U, refers to U-shaped association between UA and cognition; →, refers to no statistically significant association.

**Table S2.** Longitudinal associations of blood UA levels with the risk of MCI among participants without and with hyperuricemia.

| **Blood UA (μmol/L)** | **Model 1** | **Model 2** | **Model 3** |
| --- | --- | --- | --- |
|  | **HR (95% CI)** | **HR (95% CI)** | **HR (95% CI)** |
|  | Without hyperuricemia ^a^ | | |
| Q_1_^b^ | 1.00 [Reference] | 1.00 [Reference] | 1.00 [Reference] |
| Q_2_ | 0.73 (0.56, 0.95) | 0.73 (0.55, 0.95) | 0.74 (0.56, 0.97) |
| Q_3_ | 0.89 (0.69, 1.14) | 0.83 (0.64, 1.08) | 0.82 (0.62, 1.07) |
| Q_4_ | 0.68 (0.52, 0.90) | 0.64 (0.48, 0.86) | 0.64 (0.48, 0.85) |
|  | With hyperuricemia | | |
| Q_1_^c^ | 1.00 [Reference] | 1.00 [Reference] | 1.00 [Reference] |
| Q_2_ | 0.87 (0.38, 2.03) | 1.46 (0.58, 3.64) | 1.36 (0.50, 3.67) |
| Q_3_ | 1.16 (0.56, 2.40) | 1.29 (0.57, 2.92) | 1.23 (0.51, 2.96) |
| Q_4_ | 1.13 (0.48, 2.65) | 2.10 (0.82, 5.36) | 2.19 (0.81, 5.88) |

Abbreviations: MCI, mild cognitive impairment; UA, uric acid; HR, hazard ratio; CIs, confidence intervals.

Note: Model 1 adjusted for age, sex and education; model 2 additionally adjusted for drinking, smoking, marital status, regular exercise, body mass index, central obesity, and adequate medical service based on model 1; model3 additionally adjusted for hypertension, diabetes mellitus, self-reported history of heart disease, and stroke and cardiovascular disease based on model 2.

Q_1_ was defined as the reference group.

^a^ Serum UA levels>360 μmol/L in women and serum UA levels >420 μmol/L in men are defined as hyperuricemia.

^b^ In participants without hyperuricemia, the cutoff values of UA quartiles were 206.5, 250.3, and 293.5 µmol/L for women, and 246.7, 292.1, and 342.0 μmol/L for men.

^c^ In participants with hyperuricemia, the cutoff values of UA quartiles were 378.7, 401.6, and 449.5 µmol/L for women, and 440.5, 471.0, and 517.1 μmol/L for men.

**Table S3.** Baseline characteristics of the included and excluded study participants.

| **Characteristics** | **Participants, No. (%)** | |  |  |
| --- | --- | --- | --- | --- |
|  | **Total (n=5074)** | **Excluded (n=1971)** | **Included (n=3103)** | **P value** |
| Age, mean ± SD, years | 83.6 ± 16.0 | 81.3 ± 20.9 | 85.1 ± 11.7 | <0.001 |
| Blood UA, means ± SD | 289.7 ± 89.2 | 283.3 ± 87.4 | 293.3 ± 90.0 | 0.001 |
| Sex, women (%) | 2883 (56.9) | 1195 (60.8) | 1688 (54.4) | <0.001 |
| ≥1 Years of education, yes (%) | 1938 (38.9) | 840 (42.8) | 1098 (36.3) | <0.001 |
| Currently married, yes (%) | 1940 (38.9) | 770 (39.4) | 1170 (38.5) | 0.558 |
| Regular exercise, yes (%) | 743 (16.2) | 191 (12.1) | 552 (18.4) | <0.001 |
| Current smoking, yes (%) | 808 (17.4) | 235 (14.7) | 573 (18.8) | 0.001 |
| Current alcohol drinking, yes (%) | 755 (16.3) | 245 (15.3) | 510 (16.8) | 0.202 |
| BMI, mean ± SD, kg/m^2^ | 21.4 ± 8.4 | 21.3 ± 4.6 | 21.5 ± 10.1 | 0.375 |
| Central obesity, yes ^a^ (%) | 2045 (41.1) | 742 (39.1) | 1303 (42.3) | 0.023 |
| Adequate medical service, yes (%) | 4358 (93.7) | 1494 (93.2) | 2864 (93.9) | 0.351 |
| Hypertension, yes (%) | 2649 (52.4) | 919 (46.9) | 1730 (55.8) | <0.001 |
| Diabetes mellitus, yes (%) | 511 (10.1) | 180 (9.3) | 331 (10.7) | 0.117 |
| Heart disease, yes (%) | 329 (6.7) | 97 (5.2) | 232 (7.7) | 0.001 |
| Stroke/CVD, yes (%) | 284 (5.8) | 119 (6.3) | 165 (5.4) | 0.200 |

Abbreviations: UA, uric acid; SD, standard deviation; BMI, body mass index; CVD, cardiovascular disease.

Note: Values are given as No. (%) unless otherwise stated.

^a^ Central obesity (yes vs no) was defined as waist circumference (WC) ≥80 cm in women and WC ≥85 cm in men.

**Table S4.** Baseline characteristics of study participants by different enrolled times.

| **Characteristics** | **Total** | **2008 wave** | **2012 wave** | **2014 wave** | **P Value** |
| --- | --- | --- | --- | --- | --- |
|  | **(N=3103)** | **(N=949)** | **(N=1307)** | **(N=847)** |  |
| Age, mean ± SD, years | 85.1 ± 11.7 | 86.0 ± 12.0 | 83.5 ± 11.8 | 86.5 ± 10.7 | <0.001 |
| Blood UA, means ± SD | 293.3 ± 90.0 | 278.6 ± 88.4 | 298.2 ± 93.8 | 302.0 ± 83.7 | <0.001 |
| Sex, women (%) | 1688 (54.4) | 546 (57.5) | 669 (51.2) | 473 (55.8) | 0.007 |
| ≥1 Years of education, yes (%) | 1098 (36.3) | 297 (31.4) | 536 (41.7) | 265 (33.5) | <0.001 |
| Currently married, yes (%) | 1170 (38.5) | 310 (32.7) | 570 (45.3) | 290 (34.9) | <0.001 |
| Regular exercise, yes (%) | 552 (18.4) | 194 (20.4) | 222 (18.0) | 136 (16.4) | 0.086 |
| Current smoking, yes (%) | 2476 (81.2) | 742 (78.2) | 1012 (80.4) | 722 (85.9) | 0.000 |
| Current alcohol drinking, yes (%) | 2532 (83.2) | 771 (81.2) | 1036 (82.4) | 725 (86.7) | 0.005 |
| BMI, mean ± SD, kg/m^2^ | 21.5 ± 10.1 | 20.3 ± 3.6 | 21.7 ± 7.6 | 22.5 ± 16.7 | <0.001 |
| Central obesity, yes ^a^ (%) | 1777 (57.7) | 573 (60.6) | 766 (59.1) | 438 (52.2) | 0.001 |
| Adequate medical service, yes (%) | 2864 (93.9) | 847 (89.3) | 1195 (94.9) | 822 (97.6) | <0.001 |
| Hypertension, yes (%) | 1730 (55.8) | 422 (44.6) | 770 (59.0) | 538 (63.5) | <0.001 |
| Diabetes mellitus, yes (%) | 331 (10.7) | 110 (11.6) | 112 (8.6) | 109 (12.9) | 0.004 |
| Heart disease, yes (%) | 232 (7.7) | 58 (6.2) | 99 (7.7) | 75 (9.5) | 0.036 |
| Stroke/CVD, yes (%) | 165 (5.4) | 33 (3.5) | 86 (6.7) | 46 (5.8) | 0.005 |

Abbreviations: UA, uric acid; SD, standard deviation; BMI, body mass index; CVD, cardiovascular disease.

Note: Values are given as No. (%) unless otherwise stated. In 2008 to 2014 wave, the numbers of participants with missing data are 2, 21 and 56 in education; 35, 53 and 95 in BMI; 4, 11 and 8 in central obesity; 3, 1 and 0 in Hypertension; 110, 112 and 109 in diabetes mellitus; 7, 20 and 55 in heart disease; 6, 16 and 49 in stroke/CVD, respectively.

^a^ Central obesity (yes vs no) was defined as waist circumference (WC) ≥80 cm in women and WC≥85 cm in men.

**Table S5.** Baseline characteristics of study participants by UA quartiles for the cross-sectional analyses

| **Characteristics** | **Total** | **Q1** ^a^ | **Q2** | **Q3** | **Q4** | **P value** |
| --- | --- | --- | --- | --- | --- | --- |
|  | **(N=3899)** | **(N=975)** | **(N=972)** | **(N=976)** | **(N=976)** |  |
| Age, mean ± SD, years | 86.7 ± 12.1 | 84.9 ± 12.8 | 86.5 ± 12.0 | 87.3 ± 11.5 | 88.1 ± 11.6 | <0.001 |
| Sex, women | 2227 (57.1) | 557 (57.1) | 554 (57.0) | 558 (57.2) | 558 (57.2) | 1.000 |
| ≥1 years of education，yes | 1313 (34.3) | 321 (33.4) | 316 (33.0) | 334 (34.9) | 342 (35.9) | 0.510 |
| Currently married, yes | 1329 (34.3) | 348 (35.8) | 342 (35.5) | 333 (34.3) | 306 (31.5) | 0.181 |
| Regular exercise, yes | 634 (16.6) | 121 (12.6) | 171 (17.9) | 173 (18.1) | 169 (17.7) | 0.002 |
| Current smoking, yes | 668 (17.2) | 169 (17.4) | 155 (16.0) | 168 (17.3) | 176 (18.1) | 0.648 |
| Current alcohol drinking, yes | 603 (15.6) | 144 (14.8) | 134 (13.9) | 159 (16.4) | 166 (17.1) | 0.193 |
| BMI, mean ± SD, kg/m^2^ | 21.2 ± 9.2 | 20.6 ± 3.4 | 21.6 ± 15.4 | 21.3 ± 5.9 | 21.5 ± 7.3 | 0.083 |
| Central obesity ^b^, yes | 1530 (39.7) | 347 (36.0) | 371 (38.8) | 383 (39.7) | 429 (44.3) | 0.002 |
| Adequate medical service, yes | 3639 (93.6) | 892 (92.1) | 908 (93.8) | 923 (94.7) | 916 (93.9) | 0.112 |
| Hypertension, yes | 2180 (56.0) | 470 (48.3) | 509 (52.5) | 589 (60.3) | 612 (62.8) | <0.001 |
| Diabetes mellitus, yes | 424 (10.9) | 96 (9.8) | 83 (8.5) | 123 (12.6) | 122 (12.5) | 0.007 |
| Heart Disease, yes | 278 (7.3) | 72 (7.5) | 57 (6.0) | 72 (7.6) | 77 (8.2) | 0.333 |
| Stroke/CVD, yes | 231 (6.1) | 72 (7.5) | 50 (5.3) | 50 (5.2) | 59 (6.2) | 0.130 |

Abbreviations: UA, uric acid; BMI, body mass index; SD, standard deviation; CVD, cardiovascular disease.

Note: Values are given as No. (%) unless otherwise stated.

^a^ The cutoff values were 214.2, 264.0, and 319.0 µmol/L for women, and 252.0, 304.6, and 364.5 µmol/L for men.

^b^ Central obesity (yes vs no) was defined as waist circumference (WC) ≥80 cm in women and WC ≥85 cm in men.

**Table S****6.** Cross-sectional associations of blood UA levels with the cognitive score.

| **Blood UA (μmol/L)** | **Model 1** | **Model 2** | **Model 3** |
| --- | --- | --- | --- |
|  | **β (SE)** | **β (SE)** | **β (SE)** |
| Continuous variable |  |  |  |
| Per 10 μmol/L increment | 0.09 (0.01) ^**^ | 0.08 (0.01) ^**^ | 0.08 (0.01) ^**^ |
| By quartiles **^a^** |  |  |  |
| Q_1_ | 1.00 [Reference] | 1.00 [Reference] | 1.00 [Reference] |
| Q_2_ | 0.85 (0.35) | 0.46 (0.35) | 0.32 (0.35) |
| Q_3_ | 1.53 (0.35) ^**^ | 1.44 (0.36) ^**^ | 1.27 (0.36) ^**^ |
| Q_4_ | 1.90 (0.36) ^**^ | 1.66 (0.36) ^**^ | 1.41 (0.36) ^**^ |
| P for linear trend | <0.010 | <0.010 | <0.010 |

Abbreviations: UA, uric acid; SE, standard estimation.

Model 1 adjusted for age, sex and education; model 2 additionally adjusted for drinking, smoking, marital status, regular exercise, body mass index, central obesity, adequate medical service; model3 additionally adjusted for hypertension, diabetes mellitus, self-reported history of heart disease, and stroke and cardiovascular disease.

^a^ The cutoff values were 214.2, 264.0, and 319.0 µmol/L for women, 252.0, 304.6, and 364.5 µmol/L for men.

^**^*P*<0.01.

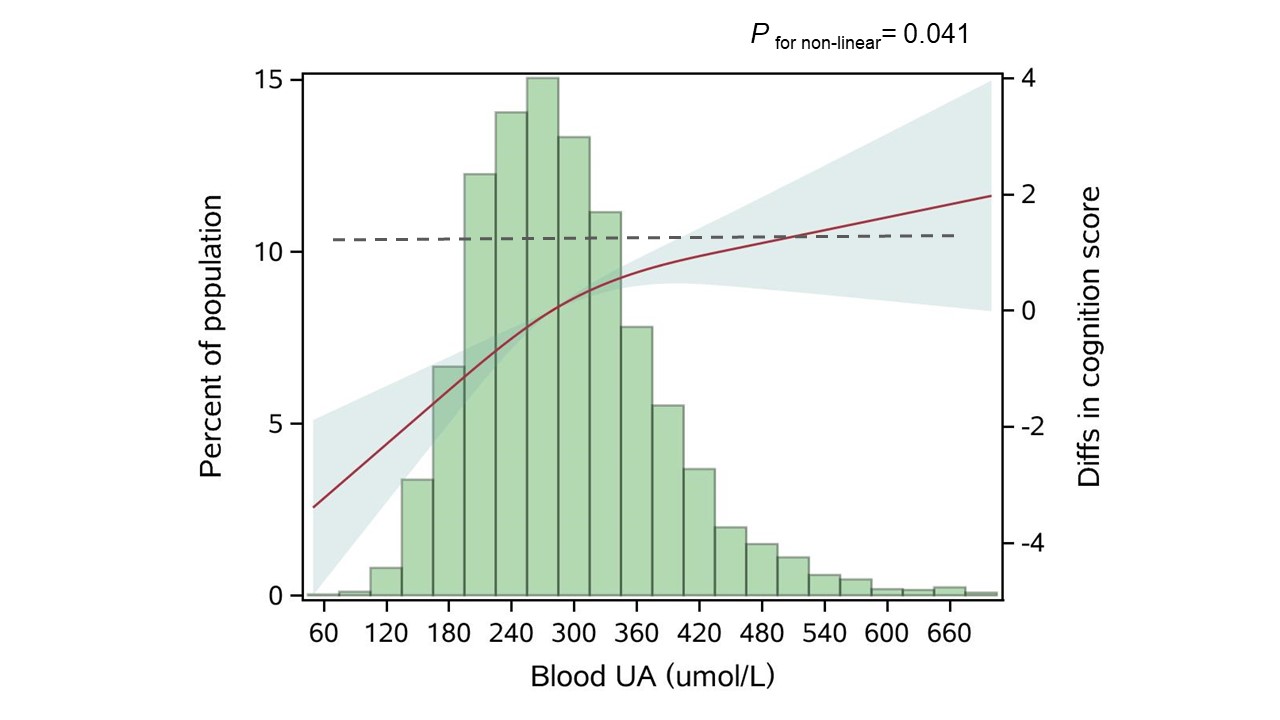

**Fig. S1.** Adjusted dose-response association between blood UA and cognition score at baseline. Abbreviation: UA, uric acid

Note: UA was coded using a restricted cubic spline (RCS) function with three sex-specific knots, which approximately corresponded to the the 5^th^ (170.5 μmol/L), 50^th^ (279.6 μmol/L), and 95^th^ (451.0 μmol/L) percentiles of blood UA distribution.

Solid red line represents the adjusted estimated coefficient for cognitive score for any value of UA compared to participants with 279.6 μmol/L (P50) of blood UA level, with light green shaded areas showing 95% confidence intervals derived from restricted cubic spline regressions. The green histograms show the fraction of the population with the different levels of blood UA. Dashed gray line refers to the reference for the association at an estimated coefficient of 0.

*P* _for non-linear_= 0.041.
